# Quantifying the impact of the cluster clinic model on geographic service accessibility

**DOI:** 10.1101/2023.12.09.23299757

**Authors:** K Johnstone, S Turner

## Abstract

**Background:** Travel distance is a barrier to accessing care, especially in large areas with low population density. The cluster clinic model is designed to provide paediatric scheduled care in the community setting within NHS Grampian. Here, we have quantified the effect on overall travel distance and equality of access.

**Methods:** Three models were compared: All clinics delivered at Royal Aberdeen Children’s Hospital, all clinics delivered at cluster clinics, and clinics at both hospital and cluster clinics. Shortest drivable distance from home to clinic in each model was calculated for all children in Aberdeen City and Aberdeenshire. Equality of distribution was assessed using a Gini coefficient, with values closer to 0 representing better equality, and 1 representing worse equality.

**Results:** In model A, median travel distance was 8.64miles (Q1: 2.96miles, Q3: 25.7miles) with a Gini coefficient of 0.491. For model B, median travel distance was 3.14miles (Q1: 1.46miles, Q3: 9.07miles) with a Gini coefficient of 0.480. In model C, median travel distance was 3.13miles (Q1: 1.35miles, Q3: 9.07miles), with a Gini coefficient of 0.490.

**Conclusions:** The cluster clinic model significantly reduces travel distance, whilst simultaneously improving access equality. This methodology should be considered to prospectively evaluate implementation of similar models elsewhere.

## Introduction

Travel distance is a fundamental barrier to accessing healthcare for many patients, with cost implications in terms of time, money, and pollution. In large geographic areas with low population density, travel distance is particularly apparent and contributes to inequity of service accessibility. NHS Grampian is one such area, with around 500,000 people (85,000 under 16) living across 3,000 square miles (Figure 1).^1^

**Figure 1:**
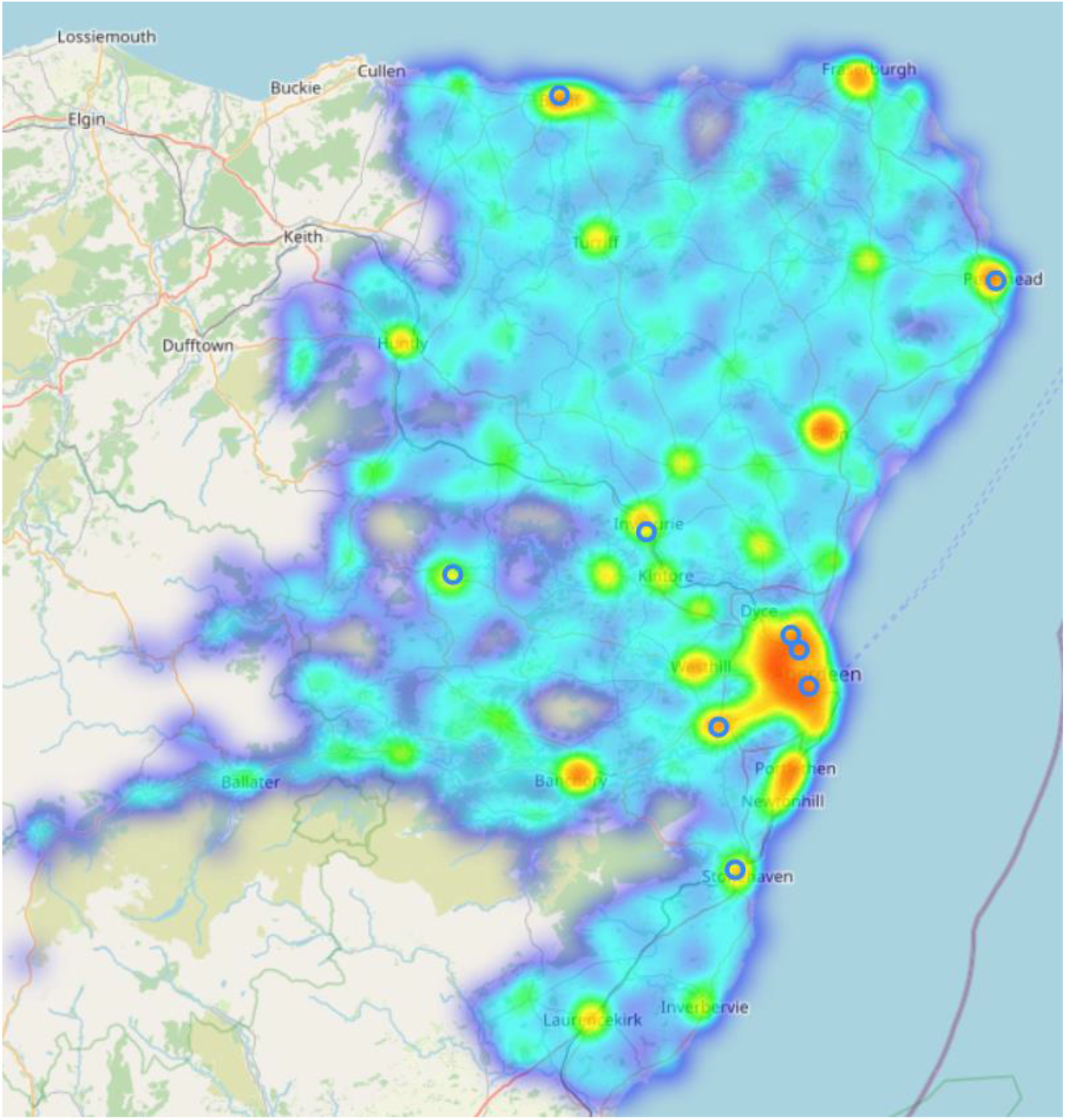
Heatmap showing population distribution in Aberdeen City and Aberdeenshire created using Centresurv, with cluster clinics shown as blue circles. Contains OpenStreetMap data © OpenStreetMap contributors, and 2011 Census data Source: Office for National Statistics © Crown Copyright 2022.

The Cluster Clinic Model is a novel approach to providing general paediatric scheduled care in a community setting in Aberdeen and Aberdeenshire. A cluster is a group of GP practices which are geographically co-located, similar to a primary care network in England.^2^ Each Cluster Clinic takes place in a community setting, and is staffed by a named paediatrician and GP with an extended role in paediatrics. Prior to the COVID-19 pandemic, cluster clinics were intended to take place in nine locations across Aberdeen City and Aberdeenshire. However, due to reduced availability of community premises, only three cluster clinics were established in community settings, with the rest being hosted at Royal Aberdeen Children’s Hospital (RACH). Prior to this model, all clinics took place at RACH.

## Methods

We compared three models of service delivery. The first (Model A) assumes delivery of all scheduled care at RACH. The second (Model B) assumes delivery of all appointments at the nine cluster clinic sites (Figure 1), and the third (Model C) proposes that the child attends either RACH or cluster clinic, whichever is the nearest.

For each postcode within the study area, the shortest drivable route to the nearest centre (RACH or cluster clinic) was calculated, using the software package centresurv.^3^ Using 2011 census data^4^, adjustments were made to account for differences in the number of people living at each postcode. Variation in distribution of children between postcodes was accounted for using data published for small data zones.^5^ This provided an accurate approximation for the travel distance of an individual child from home to clinic.

Equality of access was assessed using a Gini coefficient, in which values closer to 0 represent better equality, and values closer to 1 represent worse equality. Perfect equality (Gini=0) would reflect a system in which all patients are equidistant from their nearest centre. Conversely, as values trend towards 1, this reflects an increasingly small proportion of the population having disproportionately long travel distances relative to the overall population.

Comparison of median travel distances between travel distances was made using a Wilcoxon test, with a p value of less than 0.05 considered to be significant.

Ethics approval was not required for this study.

## Results

14,589 residential postcodes were included, with a total 84,463 children. Median travel distance for a child in Model A was 8.64miles (Q1: 2.96miles, Q3: 25.7miles). In Model B, the median distance to cluster clinic was 3.14miles (Q1: 1.46miles, Q3: 9.07miles) (Figure 2). Overall, this corresponded with shorter journeys for 74,536 children (Med: 5.49miles, Q1: 1.55miles, Q3: 15.3miles). However, 9.927 children have longer journeys (Med: 0.594miles, Q1: 0.271miles, Q3: 0.938miles). In Model C median travel distances was 3.13miles (Q1: 1.35miles, Q3: 9.07miles).

**Figure 2:**
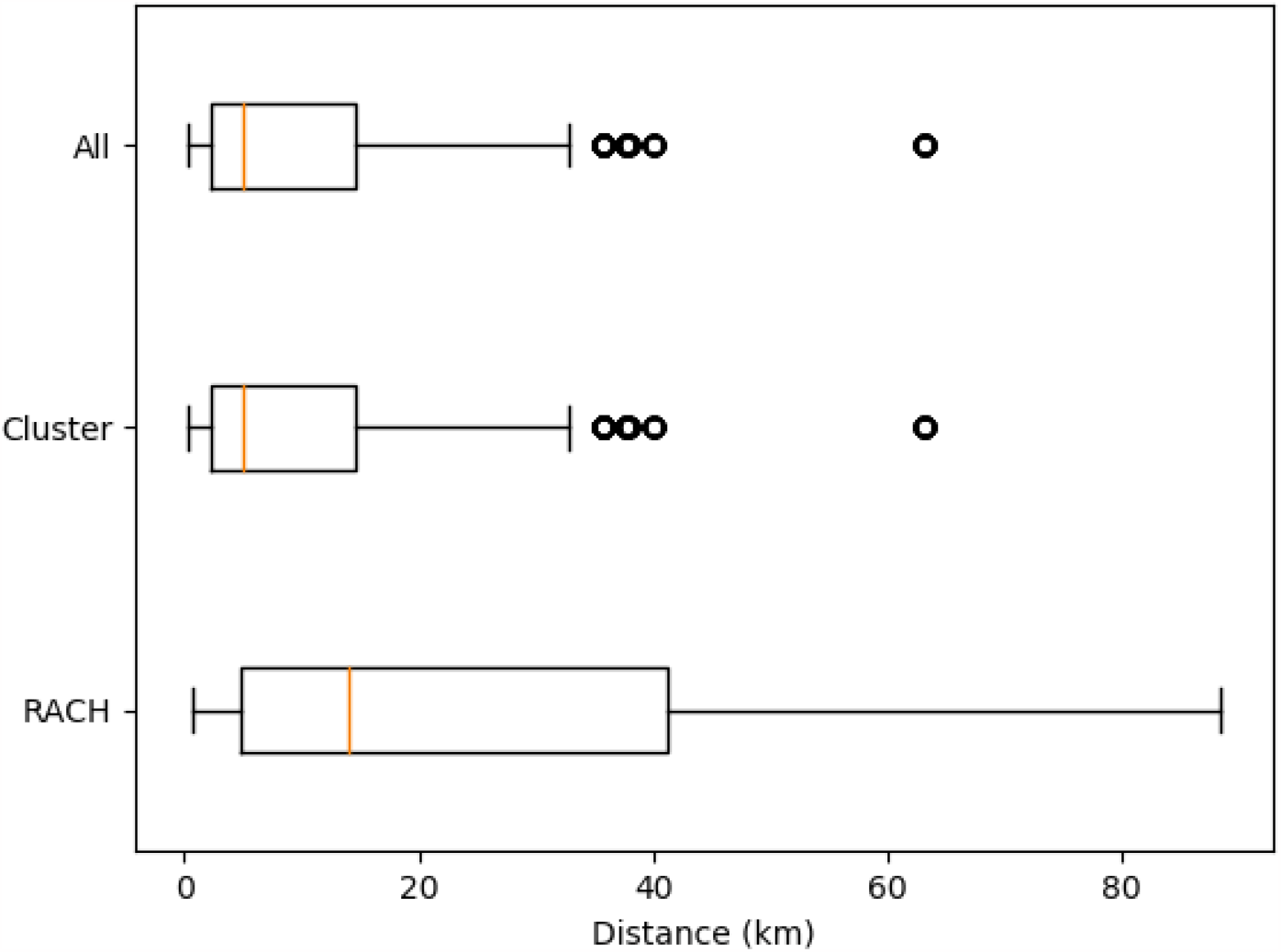
Box and whisker plot showing the distribution of travel distances to RACH (Model A), Cluster clinics (Model B), and all centres (Model C). Whiskers represent 1.5 times the interquartile range above and below Q1 and Q3. Outliers are shown as circles.

Equality of distribution was also seen to improve with a Gini coefficient of 0.491 in model A, 0.480 in model B, and 0.490 in model C.

## Discussion

The cluster clinic model was expected to reduce travel distance for patients, but this work formally quantifies the benefit, with most patients seeing travel distance half, and around a quarter of patients seeing a threefold reduction in travel distance. As such, redistributing clinics into the community significantly increases geographic accessibility, which brings many potential benefits. Shorter journeys reduce time spent travelling, with reduced carbon emissions, and make active travel options such as walking or cycling more viable.

The use of a Gini coefficient provides simple quantification of equality. Although additional centres will reduce average travel distance, if new centres are not distributed equitably throughout the population, equality of access can be made worse, with the most poorly served becoming disproportionately worse off. As such, it is reassuring to see that with just one clinic per cluster, the cluster clinic model is seen to modestly improve equality of access, reflecting proportionate improvements in access in models B and C.

Travel distance was calculated as the shortest drivable distance as this is easily measured objectively and accurately. However, this may not be the fastest or most convenient route, as speed is heavily influenced by variables such as speed limits, road conditions and traffic. Within Grampian this is most notable when considering the three small towns of Banchory, Huntly and Alford. Each of these has insufficient populations to justify a dedicated cluster clinic itself. With a cluster clinic located in Alford, travel distance may be shorter to this site for residents of Banchory and Huntly. However, travelling towards Aberdeen may be faster and more practical, due to the layout of main roads.

This analysis is a best-case analysis, which assumes that all children would attend the clinic located nearest to their home. As cluster clinic allocation is based on registered GP practice, there are likely to be a small number of cases in which the nearest clinic may not be the one to which they are allocated.

Further studies should look to establish association between clinical outcomes and journey distances.

## Conclusions

The Cluster Clinic model results in significant reductions in travel distances to clinics, whilst simultaneously improving access equality. However, careful consideration of clinic location is required to maximise benefits, ensuring the system is both efficient equitable. The methodology applied to evaluate this model can be used to simulate and optimise the application of a similar model in different contexts.

## Data Availability

All data produced in the present study are available upon reasonable request to the authors

https://gitlab.com/krj15/cluster-clinics

## Notes

### Competing Interest Statement

The authors have declared no competing interest.

### Funding Statement

This study did not receive any funding

